# Comparative dynamic aerosol efficiencies of three emergent coronaviruses and the unusual persistence of SARS-CoV-2 in aerosol suspensions

**DOI:** 10.1101/2020.04.13.20063784

**Authors:** A.C. Fears, W.B. Klimstra, P. Duprex, A. Hartman, S.C. Weaver, K.C. Plante, D. Mirchandani, J.A. Plante, P.V. Aguilar, D. Fernández, A. Nalca, A. Totura, D. Dyer, B. Kearney, M. Lackemeyer, J.K. Bohannon, R. Johnson, R.F. Garry, D.S. Reed, C.J. Roy

**Affiliations:** Tulane School of Medicine, Tulane National Primate Research Center, New Orleans, LA; Center for Vaccine Research, University of Pittsburgh, Pittsburgh, PA; World Reference Center for Emerging Viruses and Arboviruses, Institute for Human Infections and Immunity, and Department of Pathology and Center for Tropical Diseases, University of Texas Medical Branch, Galveston, TX; U.S. Army Medical Research Institute of Infectious Diseases, Fort Detrick, MD; National Institutes of Health, National Institute of Allergy and Infectious Diseases, Integrated Research Facility, Fort Detrick, MD

## Abstract

The emergent coronavirus, designated severe acute respiratory syndrome coronavirus-2 (SARS-CoV-2), is a zoonotic pathogen that has demonstrated remarkable transmissibility in the human population and is the etiological agent of a current global pandemic called COVID-19^1^. We measured the dynamic (short-term) aerosol efficiencies of SARS-CoV-2 and compared the efficiencies with two other emerging coronaviruses, SARS-CoV (emerged in 2002) and Middle Eastern respiratory syndrome CoV (MERS-CoV; emerged starting in 2012). We also quantified the long-term persistence of SARS-CoV-2 and its ability to maintain infectivity when suspended in aerosols for up to 16 hours.

---

The dynamic short-term aerosol efficiencies of SARS-CoV, SARS-CoV-2 and MERS-CoV were analyzed using three nebulizers, the Collison 3-jet, Collison 6-jet, and Aerogen Solo, to generate viral aerosols (Methods, Supplementary Appendix). Comparative efficiency experiments were performed across four separate aerobiology laboratories (Tulane University, **S1**; the National Institutes of Health Integrated Research Facility (NIH-IRF), **S2**; the United States Army Medical Institute for Infectious Diseases (USAMRIID), **S3**; and the University of Pittsburgh, **S4**). The aerosol size distributions produced by the generators used, in mass median aerodynamic diameter (MMAD), ranged from 1-3 µm with a geometric heterodispersity ranging from ≈ 1.2-1.4. Aerosols were generated into nonhuman primate head-only exposure chambers (MERS-CoV or SARS-CoV-2^2^), a customized rabbit nose-only exposure chamber (MERS-CoV), or a rodent whole-body chamber (SARS-CoV) where the overall flow was approximately 1 (Tulane University) or 0.5 (NIH-IRF, USAMRIID, University of Pittsburgh) air changes per minute. Use of these chambers and corresponding flow rates allowed us to determine the dynamic efficiencies of the virus in aerosols over a range of 30 to 60s of chamber residence time. Samples were continuously collected and integrated throughout the initiation of respective nebulizers into the chamber during aerosols varying in duration from 10-30 minutes. The short-term aerosol efficiency or spray factor (*F*_*s*_) is calculated as a unitless quotient of initial titer (PFU/liter in liquid stock) to the resultant aerosol (PFU/liter aerosol) and provides a quantitative indicator for comparing airborne fitness of viruses^3,4^. **Figure 1** shows *F*_*s*_ determinations for all three viruses when collected after <1-minute chamber residence time post-aerosolization. Comparing both MERS-CoV and SARS-CoV to SARS-CoV-2 aerosols generated with a Collison 3-jet nebulizer across three laboratories, there was a small but significant improvement in *F*_*s*_ for SARS-CoV-2 compared to both SARS-CoV (*p*=0.0254) and MERS-CoV (*p*=0.0153). Comparing nebulizers, there was an improvement in *F*_*s*_ for SARS-CoV-2 with both the Collison 6-jet (*p*=0.0066) and Aerogen Solo (*p*=0.0192) compared to the Collison 3-jet, but no difference between the Collison 6-jet and Aerogen Solo (*p*=0.4674).

**Figure 1.**
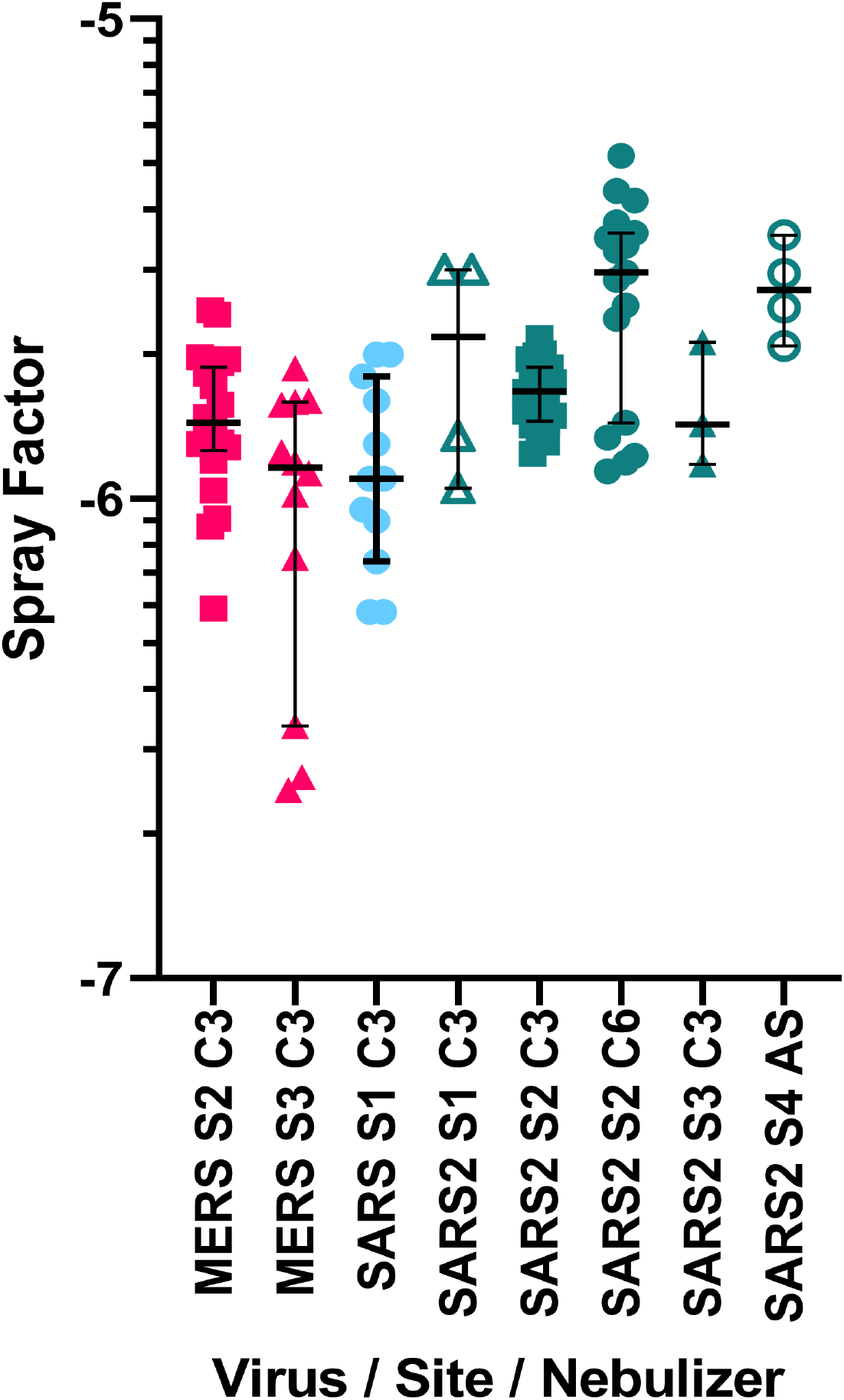
Aerosol efficiency of MERS-CoV, SARS-CoV and SARS-CoV-2 at different sites. Graph shows the spray factor (*F*_*s*_; the ratio of the nebulizer concentration to the aerosol concentration) for MERS-CoV (red symbols), SARS-CoV (blue symbols), and SARS-CoV-2 (green symbols). Aerosols were performed at four different sites (**S1** = Tulane; **S2** = NIH-IRF; **S3** = USAMRIID **S4** = Pittsburgh) and with different nebulizers (C3 = Collison 3-jet; C6 = Collison 6-jet; AS = Aerogen Solo).

Further studies with SARS-CoV-2 at one of the collaborating laboratories (Tulane University) quantified the long-term stability of airborne virus. A rotating (Goldberg) drum was used to provide an environment where the terminal settling velocity of the 2-3 µm particles is overcome by the rotational drum speed, thereby providing a static aerosol suspension of known volume^5-7^. We timed aerosol samples from the drum at 10 and 30 minutes and 2, 4, and 16 hours after initiation of rotation/suspension. The entire drum volume (10.7 liters) was evacuated at each sampling interval and represented a discrete aerosol generation event. Virus contents in collected aerosol samples were quantified by plaque assay and RT-qPCR^8^. Scanning electron microscopy was also performed on the collected aerosol samples as a qualitative assessment of virion integrity after longer-term aerosol suspension (Methods, Supplementary Appendix). Environmental parameters were measured but not controlled during the aerosol suspension experiments. The prevailing ambient environmental conditions were 23±2 °C and 53±11% relative humidity throughout the aerosol stability experiments. No UV light source was used within the cavity of the drum during suspensions. Initial titers of generated aerosols into the drum, once reaching steady-state concentration and was maintained as a static aerosol.

Infectious SARS-CoV-2 was detected at all timepoints during the aerosol suspension stability experiment (**Figure 2**). As shown in **Figure 2a**, a minor but constant fraction of the SARS-CoV-2 virus maintained replication-competence at all timepoints performed, including when sampled at 16 hours of aerosol suspension. This resulted in a remarkably flat decay curve when measured for infectivity, and failed to provide a biological half-life (κ = 2.93E-06; t_1/2_ = 2.36E+05; τ = 3.40E+05). The curve, shown in **Figure 2b** from the results of split sample analysis as quantified by qPCR, showed minimal decreases in aerosol concentration measured in viral genome copies across all of the sampled timepoints and approximated the decay curve of the infectious virus fraction shown in **Figure 2a**, including similar decay curve characteristics (κ = 6.19E-03; t_1/2_ = 111.9; τ = 161.4).

**Figure 2.**
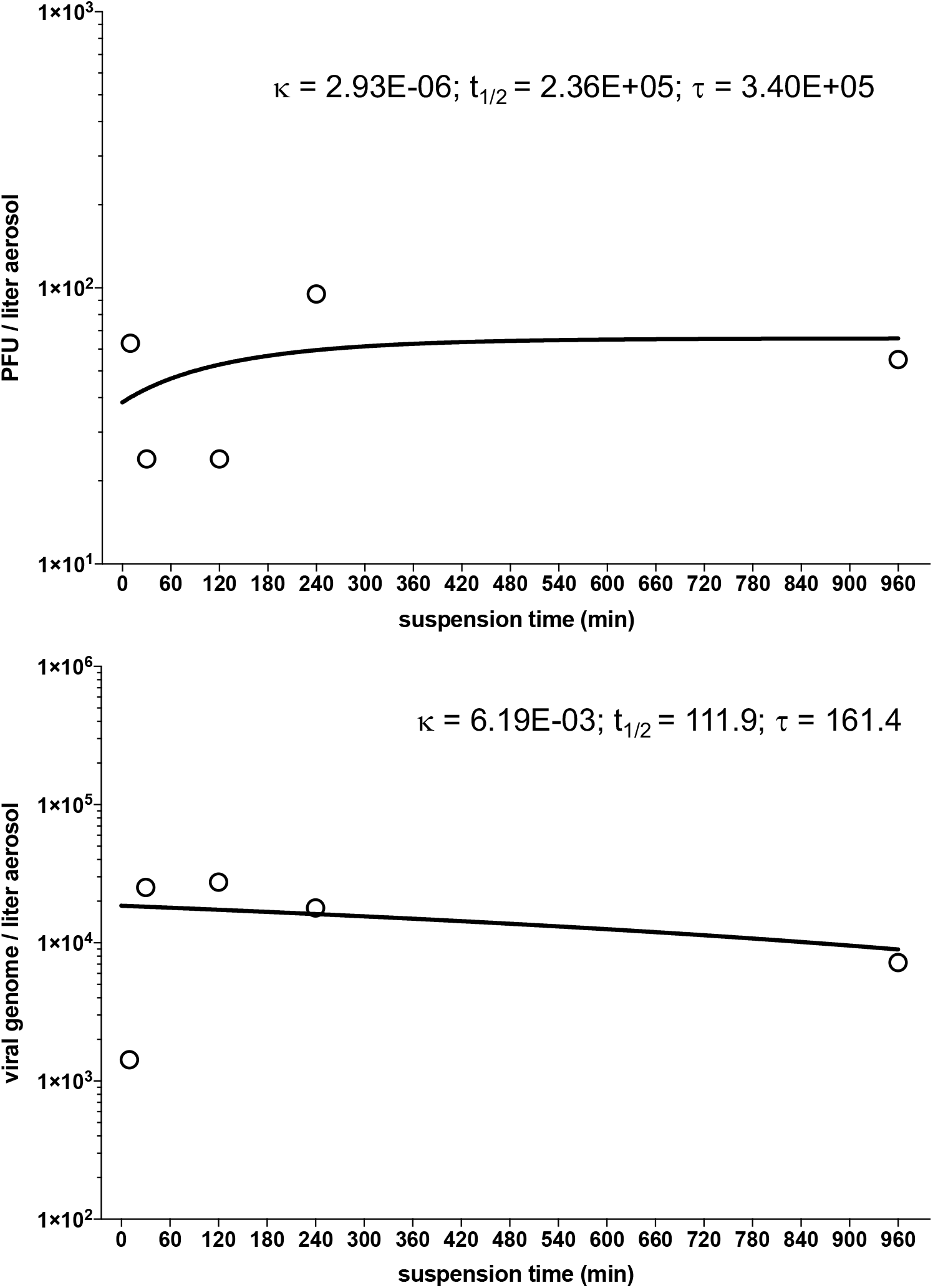
Decay curves of SARS-CoV-2 in aerosol suspension. **A**. Aerosol concentration of infectious SARS-CoV-2 as measured by plaque assay found in impinger samples collected at five differing timepoints of increased aging in aerosol suspension, **B**. Corresponding aerosol concentration of SARS-CoV-2 in time-matched impinger samples as a function of viral genome copies as measured by qPCR. Both timepoint virus estimates were graphed and nonlinear least-squares regression analysis single-order decay with no outlier detection was performed, resulting in a poor curve fit by either method of viral quantitation resulting from number and lack of iterative samples in this analysis.

We also performed a qualitative assessment of virion integrity after longer-term aerosol suspension (**Figure 3**). Scanning electron microscopy (SEM) imaging of SARS-CoV-2 revealed virions that were heterogenous in shape, either ovoid (**Figure 3A**) or spherical (**Figure 3B**). The minor:major axis ratio of oval-shaped virions was approximately 0.7, which is consistent with prior SEM analyses of SARS-CoV-2^9^. Airborne SARS-CoV-2 maintained the expected morphologies, size and aspect ratios up to 16 hours.

**Figure 3.**
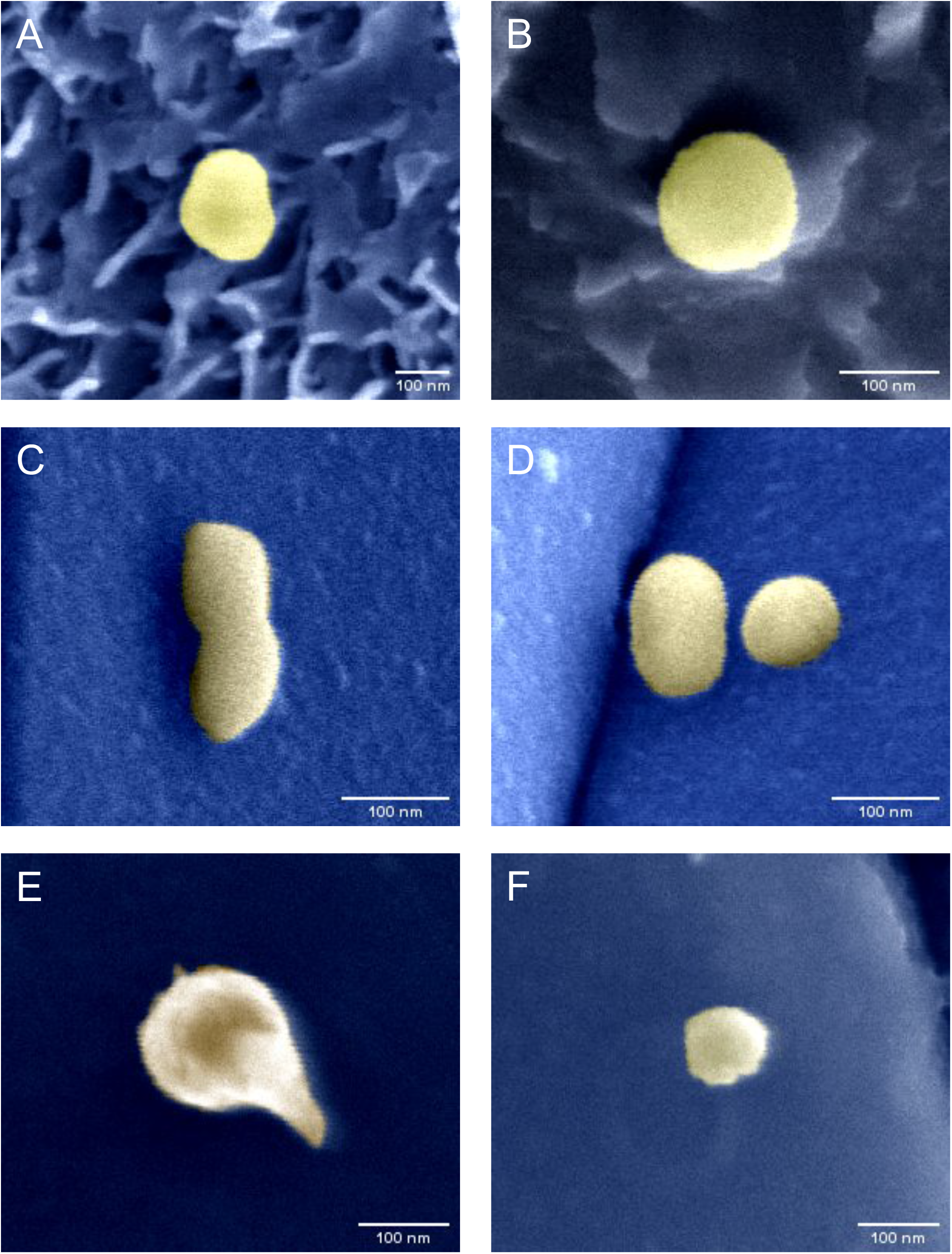
Electron microscopy images of SARS-CoV-2 in aerosol suspension at various timepoints. **A, B:** from viral stock prior to aerosolization; **C, D**: from 10 minute aerosol suspension; **E, F**: from 16 hour aerosol suspension.

Specifically, virions aged for 10 min (**Figure 3C, D**) or 16 hours (**Figure 3E, F**) were similar in shape and general appearance to virions examined prior to aerosolization, which is consistent with the retention of replication-competence.

The comparison of short-term aerosol efficiencies of three emergent coronaviruses showed SARS-CoV-2 is on par with or exceeding the efficiency estimates of SARS-CoV and MERS-CoV. Some efficiency determinations for SARS-CoV-2 ranged to -5.5^log10^, a full log difference compared to MERS-CoV. The fact that higher efficiencies trended across independent laboratories strengthens this observation. These data suggest that SARS-CoV-2 generally maintains infectivity when airborne over short distances, in contrast to either comparator betacoronavirus. Results of the aerosol suspension experiments suggest that SARS-CoV-2 is persistent over longer periods of time than would be expected when generated as a highly respirable particle (2 µm MMAD). This is remarkable, as there would be an expected decay and loss in the infectious fraction of airborne virus based on prior susceptibility studies with other relatively environmentally hardy viruses like Monkeypox^6^. A recent study^7^, showing only a slight reduction of infectivity in aerosol suspensions with approximately similar particle sizes, were suggestive of the minimal effects on SARS-CoV-2 infectivity observed in these results.

Collectively, this preliminary dataset on the aerosol efficiency and persistence of SARS-CoV-2 suggest that this virus is remarkably resilient in aerosol form, even when aged for over 12 hours, and reinforces the conclusions reached in earlier studies of aerosol fitness by others^7^. Aerosol transmission of SARS-CoV-2, whether through direct respiratory droplet transfer or fomite generation, may in fact be a more important exposure transmission pathway than previously considered^10^. Our approach of quantitative measurement of infectivity of viral airborne efficiency complemented by qualitative assessment of virion morphology leads us to conclude that SARS-CoV-2 is viable as an airborne pathogen. Humans produce aerosols continuously through normal respiration^11^. Production of aerosols increases during respiratory illnesses^12^, and even during louder-than-normal oration^13^. A fraction of naturally-generated aerosols fall within the size distribution used in our experimental studies (<5 µm), thus leading us to the conclusion that individuals infected with SARS-CoV-2 have the capacity to produce viral bioaerosols that may remain infectious over long periods of time after production via human shedding and airborne transport. Accordingly, our study results provide a basis for a broader recognition of the unique aerobiology of SARS-CoV-2, which may ultimately lead to tractable solutions and prevention interventions in the ongoing pandemic.

Alyssa C. Fears, M.S.P.H.&T.M

Robert F. Garry, Ph.D.

Chad J. Roy, Ph.D.

Tulane School of Medicine

Tulane National Primate Research Center

New Orleans, LA

Douglas S. Reed, Ph.D.

William B. Klimstra, Ph.D.

Paul Duprex, Ph.D.

Amy Hartman, Ph.D.

Center for Vaccine Research

University of Pittsburgh

Pittsburgh, PA

Scott C. Weaver, Ph.D.

Ken S. Plante, Ph.D.

Divya Mirchandani, M.S.

Jessica A. Plante, Ph.D.

World Reference Center for Emerging Viruses and Arboviruses

Institute for Human Infections and Immunity

Patricia V. Aguilar, Ph.D.

Diana Fernández, M.S.

Department of Pathology and Center for Tropical Diseases, UTMB.

University of Texas Medical Branch

Galveston, TX

Aysegul Nalca, M.D., Ph.D.

Allison Totura, Ph.D.

David Dyer,

B.S. Brian Kearney, M.S.

U.S. Army Medical Research Institute of Infectious Diseases

Fort Detrick, MD

Matthew Lackemeyer, M.S.

J. Kyle Bohannon, M.S. Reed Johnson, Ph.D. National Institutes of Health

National Institute of Allergy and Infectious Diseases Integrated Research Facility

Fort Detrick, MD

Drs. Roy and Reed contributed equally to this letter.

## Data Availability

All data can be made available for review upon request.

## Acknowledgements

The University of Pittsburgh, UTMB, and USAMRIID would like to thank Natalie Thornburg at the Centers for Disease Control for providing 2019-nCoV/USA-WA1/2020. USAMRIID thanks Dr. Kathleen Gibson for obtaining the SARS-CoV-2 virus from CDC. MERS-CoV was obtained through BEI Resources, NIAID, NIH: Middle East Respiratory Syndrome Coronavirus, EMC/2012, #NR-44260, and from the World Reference Center for Emerging Viruses and Arboviruses. The National Institute of Allergy and Infectious Diseases would like to thank Jenn Sword, Greg Kocher, and Dawn Gerhardt for providing all MERS isolates and SARS-CoV-2. The findings and conclusions in this letter are those of the authors and do not necessarily represent the official position of the United States Army or the Department of Health and Human Services. This work was supported by the Intramural Research Program of the National Institute of Allergy and Infectious Diseases, National Institutes of Health, and the the Office of the Chancellor at the University of Pittsburgh. Work performed at Tulane National Primate Research Center (CJR, ACF) was supported in part by the National Institutes of Health grant P51OD011104. RFG was supported by the National Institutes of Health grant number U19AI135995. UTMB work was supported by NIH grant R24AI120942. The Defense Health Program provided the funding for SARS-CoV-2 work in USAMRIID. The work at NIH-IRF was funded in part through the NIAID, Division of Intramural Research and Division of Clinical Research, Battelle Memorial Institute’s prime contract with NIAID under HHSN272200700016I and in whole or in part with Federal funds from the NIAID, NIH, DHHS, HHSN272201800013C. JKB performed this work as an employee of Battelle Memorial Institute. MGL performed this work as an employee of Lovelace Respiratory Research Institute and Laulima Government Solutions, LLC.

